# Blood Pressure Control and Maintenance in a Prospective Cohort of Younger Veterans: Roles of Sex, Race, Ethnicity, and Social Determinants of Health

**DOI:** 10.1101/2024.04.22.24306203

**Authors:** Allison E. Gaffey, Tiffany E. Chang, Cynthia A. Brandt, Sally G. Haskell, Sanket S. Dhruva, Lori A. Bastian, Allison Levine, Skanderson Melissa, Matthew M. Burg

## Abstract

**Background:** Proactive blood pressure (BP) management is particularly beneficial for younger Veterans, who have a greater prevalence and earlier onset of cardiovascular disease than non-Veterans. It is unknown what proportion of younger Veterans achieve and maintain BP control after hypertension onset and if BP control differs by demographics and social deprivation.

**Methods:** Electronic health records were merged from Veterans who enrolled in VA care 10/1/2001-9/30/2017 and met criteria for hypertension – first diagnosis or antihypertensive fill. BP control (140/90 mmHg), was estimated 1, 2, and 5 years post-hypertension documentation, and characterized by sex, race, and ethnicity. Adjusted logistic regressions assessed likelihood of BP control by these demographics and with the Social Deprivation Index (SDI).

**Results:** Overall, 17% patients met criteria for hypertension (n=198,367; 11% of women, median age 41). One year later, 59% of men and 65% of women achieved BP control. After adjustment, women had a 72% greater odds of BP control than men, with minimal change over 5 years. Black adults had a 22% lower odds of BP control than White adults. SDI did not significantly change these results.

**Conclusions:** In the largest study of hypertension in younger Veterans, 41% of men and 35% of women did not have BP control after 1 year, and BP control was consistently better for women through 5 years. Thus, the first year of hypertension management portends future, long-term BP control. As social deprivation did not affect BP control, the VA system may protect against disadvantages observed in the general U.S. population.

## INTRODUCTION

Men and women U.S. military Veterans show an earlier elevation in blood pressure (BP) and a greater risk of hypertension and cardiovascular disease (CVD) than non-Veterans.^1–5^ Proactive, guideline-based BP management can mitigate the proximal impact of hypertension on cardiovascular health and later CVD risk,^6^ which could be particularly beneficial for younger Veterans – i.e., those discharged from military service post-9/11.^7^ In recent years BP control has decreased in the U.S.,^8^ a decline observed across all demographic groups.^9^ Over half of younger Veterans seek care through the Department of Veterans Affairs (VA),^10^ yet it is unknown how well BP is controlled among this group. As younger adults show lower hypertension awareness, treatment and control than older adults,^11^ information concerning BP control in this at-risk, younger group of Veterans can dually inform hypertension management and improve CVD prevention.

The population of younger, post-9/11 Veterans is more demographically heterogeneous compared to those who served in earlier conflicts.^12,13^ By 2042, women will represent 20% of the Veteran population,^10^ and are more likely to use VA benefits than men.^10^ Furthermore, almost half of women Veterans are non-White.^14^ In the general population, non-Hispanic Black men and women show the greatest prevalence of hypertension.^6,15^ There are also notable variations in hypertension management and BP control by sex, race, and ethnicity.^16^ Given the high representation of women and members of racial and ethnic minorities, determining if BP control varies by demographic factors is necessary to support the cardiovascular healthcare needs of younger Veterans.

Social determinants of health (SDoH) – the conditions in which people are born, grow, work, live, and age – contribute to cardiovascular health, risk for hypertension, and CVD outcomes,^17,18^ and may underlie intersectional differences in BP control.^19^ In the general population, those with a lower income, who live in under resourced neighborhoods, and who have less healthcare access, are less likely to have controlled BP.^20^ As the VA system of care is designed to offset the adverse effects of many individual and systemic SDoH,^21^ those who receive care through the VA may experience a lower burden of social deprivation on BP management. With increasing recognition of the importance of SDoH, it is informative to explore if SDOH affect potential demographic differences in BP control.

This investigation used data from a nationwide, prospective cohort study of >1 million young and middle-aged Veterans who served in support of U.S. conflicts in Iraq and Afghanistan (post-9/11) and who enrolled in VA care to pursue four objectives. First, to determine the proportion of men and women who met criteria for hypertension based on a diagnosis or receipt of an antihypertensive medication (AHM). We hypothesized that women, particularly members of racial and ethnic minority groups, would be less likely to have a diagnosis of hypertension and AHM than their male counterparts. A second objective was to determine the proportion of patients who achieved BP control at 1, 2, and 5 years after the first evidence of hypertension. We hypothesized that rates of BP control would increase over time. A third objective was to determine if there are differences BP control based on sex, race, and ethnicity. It was hypothesized that a greater percentage of women would show BP control than men, and that White, non-Hispanic individuals would show better BP control than other racial and ethnic groups. The fourth, exploratory objective was to test if including a composite measure of SDoH would affect potential subgroup differences in BP control.

## METHODS

### Data Sources

Data used for these analyses are covered under a proprietary data use agreement with the Department of Veterans Affairs and are not available for distribution by the authors. Eligible men and women were identified from rosters maintained by the VA’s Defense Manpower Data Center – Contingency Tracking System. We extracted patients’ electronic health record (EHR) data from the national VA Corporate Data Warehouse. These data included vital sign and coded diagnostic and procedural data from all VA healthcare visits. The records were combined with additional administrative, pharmacy, and diagnostic data (based on International Classification of Diseases, Ninth Revision, Clinical Modification and Tenth Revision [ICD-9-CM and 10] codes and dates).

### Study Population

The Women Veterans Cohort Study (WVCS) is a prospective investigation of all post-9/11 men and women Veterans (i.e., those who served during conflicts in Afghanistan and Iraq, including Operations Enduring Freedom, Iraqi Freedom, and New Dawn), who were discharged from service as of October 1, 2001-September 30, 2017, and whose first VA outpatient medical visit occurred after their last date of service and before September 30, 2017. The original cohort was restricted to those with a diagnosis of hypertension, defined as i) an ICD-9 or 10 code recorded during at least 2 outpatient or 1 inpatient encounter, or; ii) at least 1 AHM prescription filled at a VA Pharmacy. In addition to meeting criteria for hypertension, eligible patients were required to have at least two qualifying BP measurements taken after their first date of VA care and before the end of the 5-year follow-up period. Furthermore, to explore potential differences in BP control by SDoH categories, patients were excluded from the analysis if they had a missing or invalid 5-digit residential zip code, or if their zip code corresponded to Puerto Rico, since linked SDoH data was unavailable (n=7,311). These criteria resulted in a final analytic sample of 198,367 men and women.

### Primary and Secondary Predictors: Sex, Race, and Ethnicity

Patients’ sex (men, women), race and ethnicity (non-Hispanic White, non-Hispanic Black, Hispanic, Asian, Other race) were extracted from the Department of Defense roster files. Sex was the primary predictor, while race and ethnicity data were combined as the secondary predictor.

### Exploratory Predictor: Social Deprivation Index

An index of SDoH was used to explore if inclusion influenced associations between the demographics and likelihood of BP control. The Social Deprivation Index (SDI) is a composite measure of a geographic area’s estimated deprivation level that is derived from 7 measures in the American Community Survey: the percentage of adults living in poverty; with <12 years education; residing in a single-parent household, a rented housing unit, or an overcrowded housing unit; households without a car; and unemployed individuals aged <65 years.^22^ Analyses were based on publicly available 2015 data (https://www.graham-center.org/maps-data-tools/social-deprivation-index.html). Area-level SDI scores were linked with patients’ residential zip codes. Scores range from 1 to 100, with higher scores indicating greater disadvantage.

### Outcome: BP control

BP control was assessed 1, 2, and 5 years after a patient first met criteria for hypertension. BP results were limited to outpatient assessments to avoid potential transient changes from acute hospitalizations.^23^ Qualifying BP measurements included those within biologically plausible ranges (systolic 70-240 mmHg; diastolic 35-140 mmHg^24^). To address multiple BP measurements within a 5-minute period: a) if measurements were identical (i.e., duplicate systolic or diastolic values), then one set was deleted; b) if there were 2 unique measurements within a 5-minute period, the means of each systolic and diastolic measurements were used to represent a single BP measurement; c) if there were >2 unique measurements within one 5-minute time window, the first BP measurement was dropped and the mean of the remaining values was used.^23^ BP control was assessed using the last 2 outpatient clinic BP measurements within 1, 2, and 5 years of the qualifying hypertension diagnosis date, and was defined by the last 2 systolic and diastolic measurements. As the duration of follow-up was generally before 2017 updates to the clinical practice guidelines for BP management,^25,26^ BP control was evaluated using a threshold of <140/90 mmHg.

### Covariates

Covariates were chosen *a priori* and measured at baseline or during the first evidence of diagnosis across follow-up. These included demographics: age (<40 vs ≥40 years) and marital status (married vs not married); behavioral/lifestyle factors: smoking status (current, past, never), body mass index (BMI; categories of underweight/normal, overweight, and obese [reference]), and history of an alcohol or substance use disorder (ever diagnosed vs. never [reference]); history of MST^27^ and other clinical risk factors (ever diagnosed vs. never [reference]): posttraumatic stress disorder (PTSD), obstructive sleep apnea (OSA), Type 1 or 2 diabetes, dyslipidemia; and the number of comorbidities using a modified version of the Charlson Comorbidity Index (i.e., this version did not include leukemia or lymphoma in the weights).^28^ Lastly, a healthcare utilization variable was created based on the number of primary care visits completed within first 2 years.^29^ Patients were also characterized by education, rurality of residence, the availability of additional, non-VA health insurance at baseline, BP category at baseline (<120/80 mmHg, 120-129/<80 mmHg, 130-139/80-89 mmHg, 140-159/90-99 mmHg, or ≥160/100 mmHg^26,30^), and if available, by the first AHM prescribed. Finally, SDI quartiles were included (ranging from Quartile 1=least disadvantaged to Quartile 4=most disadvantaged). As education and rurality are included in the calculation of SDI composites, these variables were not examined as independent covariates.

### Statistical analysis

Statistical analyses were conducted using SAS version 9.4 (Cary, NC) with *p*<0.05 as the statistical significance threshold. First, descriptive and bivariate statistics were used to summarize the data and to determine differences in demographic and clinical factors with BP control at 1, 2, and 5 years. These tests were conducted by sex, race, and ethnicity subgroups. Next, random intercept, multilevel logistic regression models were conducted using the GLIMMIX procedure to assess the associations between sex, race and ethnicity, and BP control. The odds of BP control were modeled separately for 1, 2, and 5 years. Since the primary predictors were sex, race, and ethnicity, the initial, unadjusted models only included the independent associations of sex or race/ethnicity on BP control (Model 1). Next, the logistic regression models included stepwise adjustment for: other demographics (Model 2), behavioral/lifestyle factors (Model 3), and clinical risk factors (Model 4). In additional models, we tested the two-way interactions of sex and race on BP control. Finally, exploratory analyses were also conducted to include SDI.

## RESULTS

Demographic data were available from 1,177,204 patients (12% of whom were women [n=146,091] Figure 1). Among that group, 922,543 patients (13% women [n=115,402]) had at least 1 BP measurement in an outpatient setting. As seen in Table 1, 198,367 unique patients met at least one criterion for hypertension – a diagnosis or AHM fill (11% of whom were women [n=20,814]). Of that group, 50% of patients had both a hypertension diagnosis and an AHM fill, 18% had only a diagnosis, and 33% had a prescription for an AHM but no diagnosis of hypertension. Of those who met criteria for hypertension, more men than women had both documentation of a hypertension diagnosis and an AHM prescription (51% vs. 38%). Among men and women, those who identified as non-Hispanic Black were more likely to have a hypertension diagnosis and an associated AHM fill (61% and 49%) than same-sex men and women in other subgroups.

**Figure 1.**
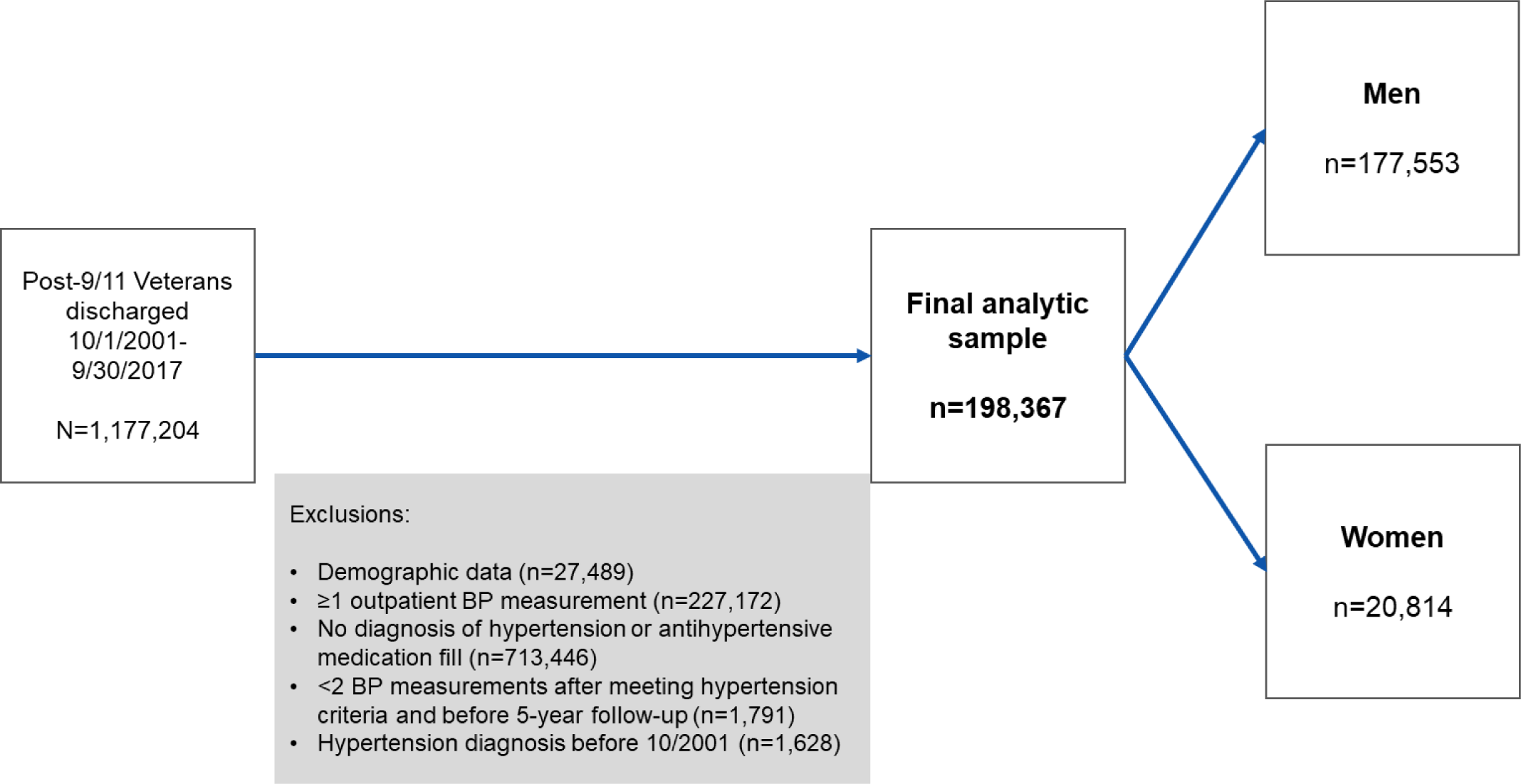
Study flowchart of the cohort of post-9/11 men and women Veterans.

**Table 1.** Percentage of patients who met criteria for hypertension, by sex, race, and ethnicity group (n=198,367).

|  |  | Men (*n*=177,553) |  |  |  |  |  | Women (*n*=20,814) |  |  |  |  |  |
| --- | --- | --- | --- | --- | --- | --- | --- | --- | --- | --- | --- | --- | --- |
|  | Total | Total | NH White | NH Black | Hispanic | NH Asian | NH Other | Total | NH White | NH Black | Hispanic | NH Asian | NH Other |
| AHM fill only | 64,873<br>(32.7) | 54,770<br>(30.9) | 37,917<br>(34.0) | 7,935<br>(20.7) | 5,948<br>(34.3) | 1,110<br>(27.2) | 1,860<br>(29.8) | 10,103<br>(48.5) | 5,347<br>(59.5) | 3,044<br>(34.4) | 1,057<br>(60.9) | 203<br>(49.3) | 452<br>(55.0) |
| Hypertension diagnosis code only | 34,608<br>(17.5) | 31,784<br>(17.9) | 19,907<br>(17.9) | 7,193<br>(18.7) | 2,890<br>(16.7) | 710<br>(17.4) | 1,084<br>(17.4) | 2,824<br>(13.6) | 989<br>(11.0) | 1,511<br>(17.1) | 181<br>(10.4) | 58<br>(14.1) | 85<br>(10.3) |
| AHM and hypertension diagnosis | 98,886<br>(49.9) | 90,999<br>(51.3) | 53,663<br>(48.1) | 23,293<br>(60.6) | 8,484<br>(49.0) | 2,256<br>(55.4) | 3,303<br>(52.9) | 7,887<br>(37.9) | 256<br>(29.5) | 4,297<br>(48.5) | 498<br>(28.7) | 151<br>(3.7) | 285<br>(34.7) |

Abbreviations: AHM, antihypertensive medication; HTN, hypertension; NH, non-Hispanic

Table 2 presents demographic, behavioral, and clinical characteristics of patients with hypertension, by sex. At baseline, the median ages of men and women were 41 and 42 years. Overall, a significantly greater percentage of women were non-White, unmarried, and lived in an urban locale compared to men. When examining behavioral factors, fewer women than men had ever smoked, were categorized as obese, or ever had a diagnosis of alcohol use disorder or substance use disorder. In relation to men, a greater percentage of women had a documented history of MST, but significantly fewer women than men had a diagnosis of PTSD, diabetes, dyslipidemia, or OSA. Women had slightly fewer AHM fills across the duration of follow-up and attended significantly more primary care visits compared to men. The average follow-up time was similar by sex – over 3 years for each group. Slightly more women vs men were in the most disadvantaged SDI quartile (29% vs 25%).

**Table 2.**
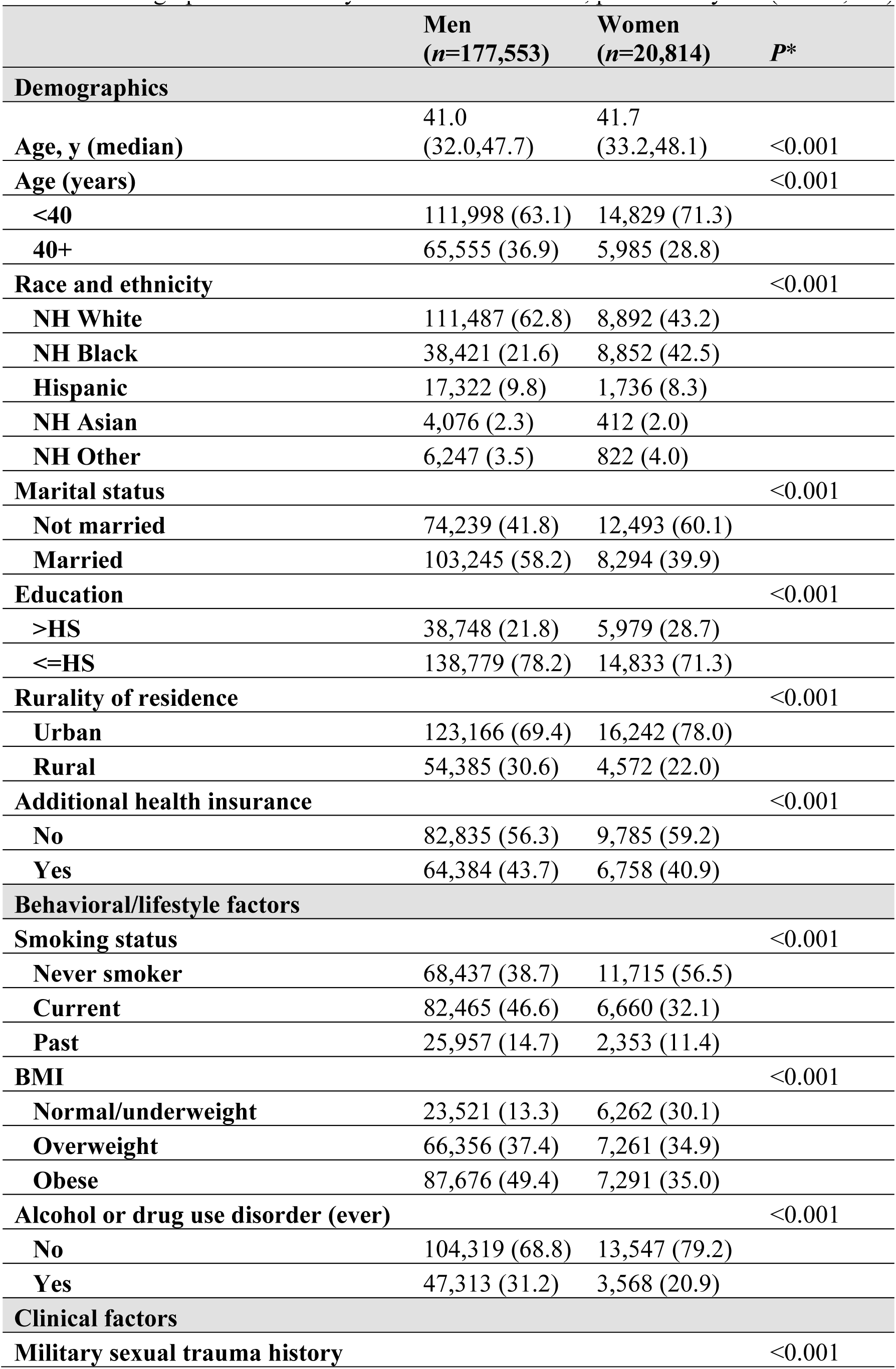
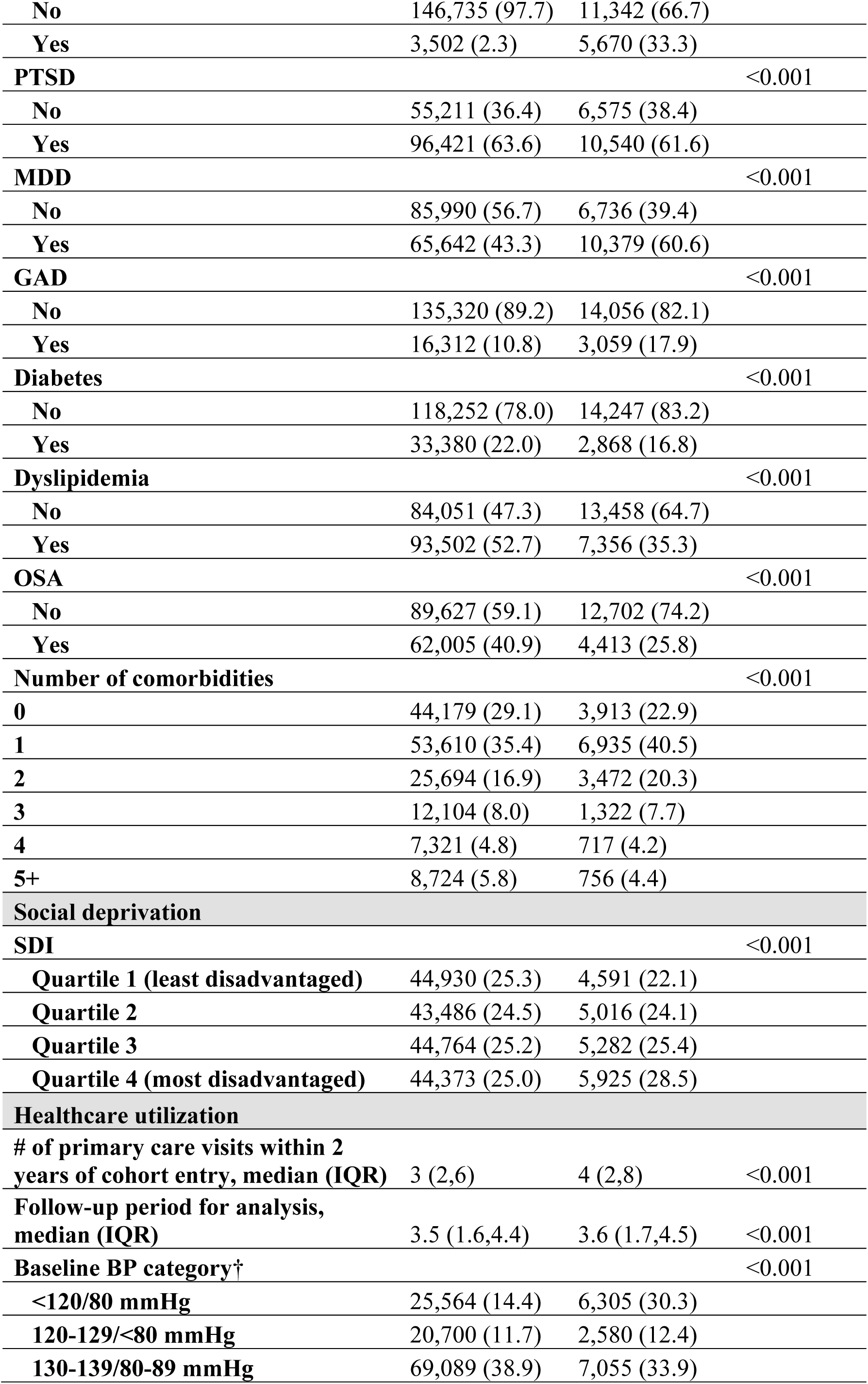
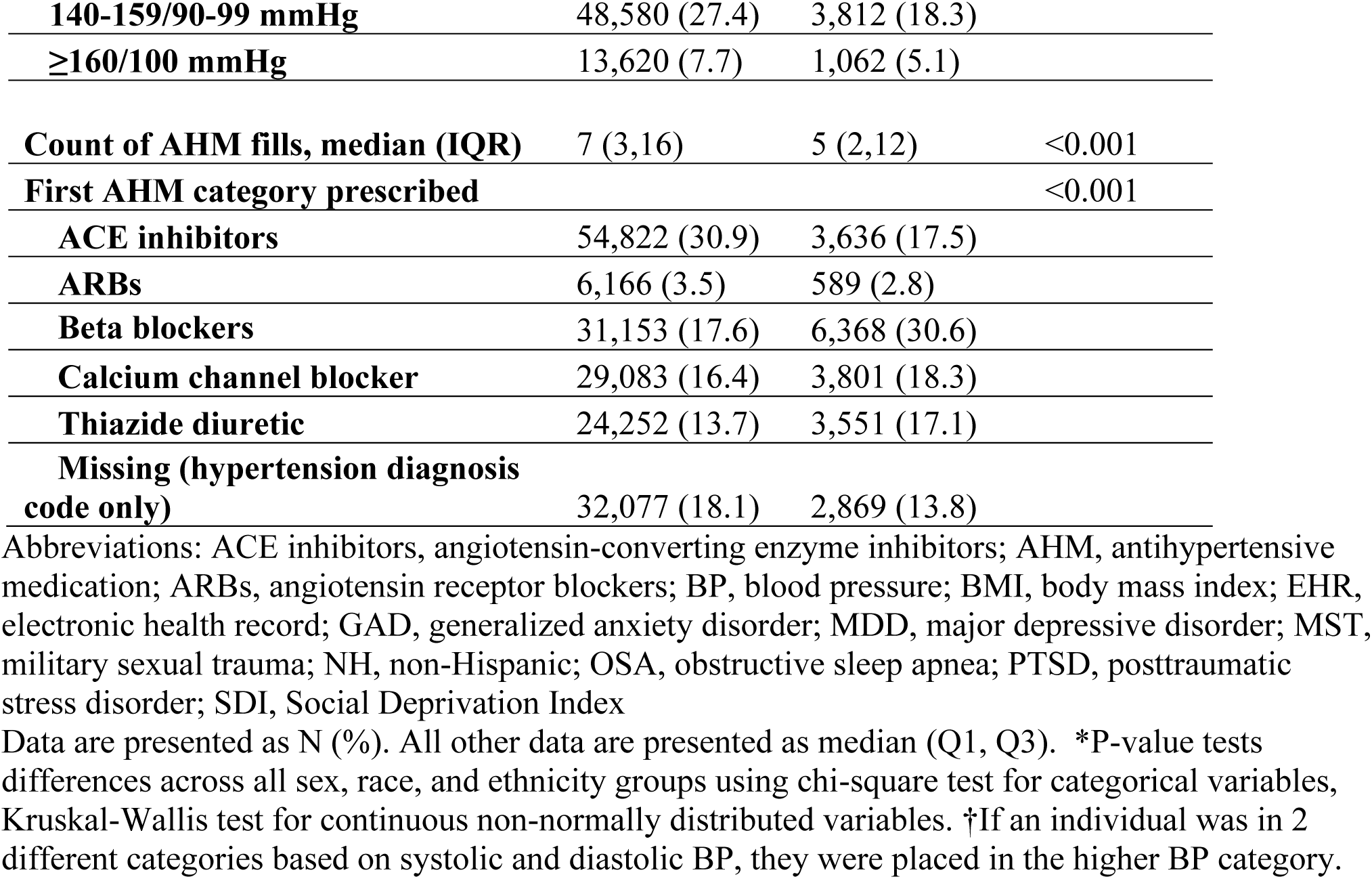
Demographic and military service characteristics, presented by sex (n=198,367).

|  | **Men**<br>**(n=177,553)** | **Women**<br>**(n=20,814)** | **P\*** |
| --- | --- | --- | --- |
| **Demographics** |  |  |  |
| **Age, y (median)** | 41.0<br>(32.0,47.7) | 41.7<br>(33.2,48.1) | <0.001 |
| **Age (years)** |  |  | <0.001 |
| <40 | 111,998 (63.1) | 14,829 (71.3) |  |
| 40+ | 65,555 (36.9) | 5,985 (28.8) |  |
| **Race and ethnicity** |  |  | <0.001 |
| NH White | 111,487 (62.8) | 8,892 (43.2) |  |
| NH Black | 38,421 (21.6) | 8,852 (42.5) |  |
| Hispanic | 17,322 (9.8) | 1,736 (8.3) |  |
| NH Asian | 4,076 (2.3) | 412 (2.0) |  |
| NH Other | 6,247 (3.5) | 822 (4.0) |  |
| **Marital status** |  |  | <0.001 |
| Not married | 74,239 (41.8) | 12,493 (60.1) |  |
| Married | 103,245 (58.2) | 8,294 (39.9) |  |
| **Education** |  |  | <0.001 |
| >HS | 38,748 (21.8) | 5,979 (28.7) |  |
| <=HS | 138,779 (78.2) | 14,833 (71.3) |  |
| **Rurality of residence** |  |  | <0.001 |
| Urban | 123,166 (69.4) | 16,242 (78.0) |  |
| Rural | 54,385 (30.6) | 4,572 (22.0) |  |
| **Additional health insurance** |  |  | <0.001 |
| No | 82,835 (56.3) | 9,785 (59.2) |  |
| Yes | 64,384 (43.7) | 6,758 (40.9) |  |
| **Behavioral/lifestyle factors** |  |  |  |
| **Smoking status** |  |  | <0.001 |
| Never smoker | 68,437 (38.7) | 11,715 (56.5) |  |
| Current | 82,465 (46.6) | 6,660 (32.1) |  |
| Past | 25,957 (14.7) | 2,353 (11.4) |  |
| **BMI** |  |  | <0.001 |
| Normal/underweight | 23,521 (13.3) | 6,262 (30.1) |  |
| Overweight | 66,356 (37.4) | 7,261 (34.9) |  |
| Obese | 87,676 (49.4) | 7,291 (35.0) |  |
| **Alcohol or drug use disorder (ever)** |  |  | <0.001 |
| No | 104,319 (68.8) | 13,547 (79.2) |  |
| Yes | 47,313 (31.2) | 3,568 (20.9) |  |
| **Clinical factors** |  |  |  |
| **Military sexual trauma history** |  |  | <0.001 |
| No | 146,735 (97.7) | 11,342 (66.7) |  |
| Yes | 3,502 (2.3) | 5,670 (33.3) |  |
| **PTSD** |  |  | <0.001 |
| No | 55,211 (36.4) | 6,575 (38.4) |  |
| Yes | 96,421 (63.6) | 10,540 (61.6) |  |
| **MDD** |  |  | <0.001 |
| No | 85,990 (56.7) | 6,736 (39.4) |  |
| Yes | 65,642 (43.3) | 10,379 (60.6) |  |
| **GAD** |  |  | <0.001 |
| No | 135,320 (89.2) | 14,056 (82.1) |  |
| Yes | 16,312 (10.8) | 3,059 (17.9) |  |
| **Diabetes** |  |  | <0.001 |
| No | 118,252 (78.0) | 14,247 (83.2) |  |
| Yes | 33,380 (22.0) | 2,868 (16.8) |  |
| **Dyslipidemia** |  |  | <0.001 |
| No | 84,051 (47.3) | 13,458 (64.7) |  |
| Yes | 93,502 (52.7) | 7,356 (35.3) |  |
| **OSA** |  |  | <0.001 |
| No | 89,627 (59.1) | 12,702 (74.2) |  |
| Yes | 62,005 (40.9) | 4,413 (25.8) |  |
| **Number of comorbidities** |  |  | <0.001 |
| 0 | 44,179 (29.1) | 3,913 (22.9) |  |
| 1 | 53,610 (35.4) | 6,935 (40.5) |  |
| 2 | 25,694 (16.9) | 3,472 (20.3) |  |
| 3 | 12,104 (8.0) | 1,322 (7.7) |  |
| 4 | 7,321 (4.8) | 717 (4.2) |  |
| 5+ | 8,724 (5.8) | 756 (4.4) |  |
| **Social deprivation** |  |  |  |
| **SDI** |  |  | <0.001 |
| Quartile 1 (least disadvantaged) | 44,930 (25.3) | 4,591 (22.1) |  |
| Quartile 2 | 43,486 (24.5) | 5,016 (24.1) |  |
| Quartile 3 | 44,764 (25.2) | 5,282 (25.4) |  |
| Quartile 4 (most disadvantaged) | 44,373 (25.0) | 5,925 (28.5) |  |
| **Healthcare utilization** |  |  |  |
| # of primary care visits within 2 years of cohort entry, median (IQR) | 3 (2,6) | 4 (2,8) | <0.001 |
| Follow-up period for analysis, median (IQR) | 3.5 (1.6,4.4) | 3.6 (1.7,4.5) | <0.001 |
| Baseline BP category† |  |  | <0.001 |
| <120/80 mmHg | 25,564 (14.4) | 6,305 (30.3) |  |
| 120-129/<80 mmHg | 20,700 (11.7) | 2,580 (12.4) |  |
| 130-139/80-89 mmHg | 69,089 (38.9) | 7,055 (33.9) |  |
| **140-159/90-99 mmHg** | 48,580 (27.4) | 3,812 (18.3) |  |
| **≥160/100 mmHg** | 13,620 (7.7) | 1,062 (5.1) |  |
| **Count of AHM fills, median (IQR)** | 7 (3,16) | 5 (2,12) | <0.001 |
| **First AHM category prescribed** |  |  | <0.001 |
| **ACE inhibitors** | 54,822 (30.9) | 3,636 (17.5) |  |
| **ARBs** | 6,166 (3.5) | 589 (2.8) |  |
| **Beta blockers** | 31,153 (17.6) | 6,368 (30.6) |  |
| **Calcium channel blocker** | 29,083 (16.4) | 3,801 (18.3) |  |
| **Thiazide diuretic** | 24,252 (13.7) | 3,551 (17.1) |  |
| **Missing (hypertension diagnosis code only)** | 32,077 (18.1) | 2,869 (13.8) |  |

Abbreviations: ACE inhibitors, angiotensin-converting enzyme inhibitors; AHM, antihypertensive medication; ARBs, angiotensin receptor blockers; BP, blood pressure; BMI, body mass index; EHR, electronic health record; GAD, generalized anxiety disorder; MDD, major depressive disorder; MST, military sexual trauma; NH, non-Hispanic; OSA, obstructive sleep apnea; PTSD, posttraumatic stress disorder; SDI, Social Deprivation Index
Data are presented as N (%). All other data are presented as median (Q1, Q3). \*P-value tests differences across all sex, race, and ethnicity groups using chi-square test for categorical variables, Kruskal-Wallis test for continuous non-normally distributed variables. †If an individual was in 2 different categories based on systolic and diastolic BP, they were placed in the higher BP category.

Table 3 presents characteristics of men and women who met criteria for hypertension by sex, race, and ethnicity. Of the men who met criteria for hypertension, White, Hispanic, and Other-identified groups had the highest percentage of individuals who were under age 40 and unmarried. In all racial and ethnic groups, more men lived in urban vs. rural settings. Over half of White, non-Hispanic men were current smokers, which was a significantly greater prevalence than observed in other groups. White men were less likely to have a diagnosis of diabetes or OSA compared to the other groups. Hispanic men showed the greatest prevalence of PTSD, depression, and OSA compared to others. Based on BP assessed at baseline, the highest percentage of men in each race and ethnicity group were categorized as 130-139/80-80mmHg (37-40%) and each race and ethnicity group were similarly prescribed an angiotensin converting enzyme inhibitor (ACE-inhibitor) as their first AHM.

**Table 3.**
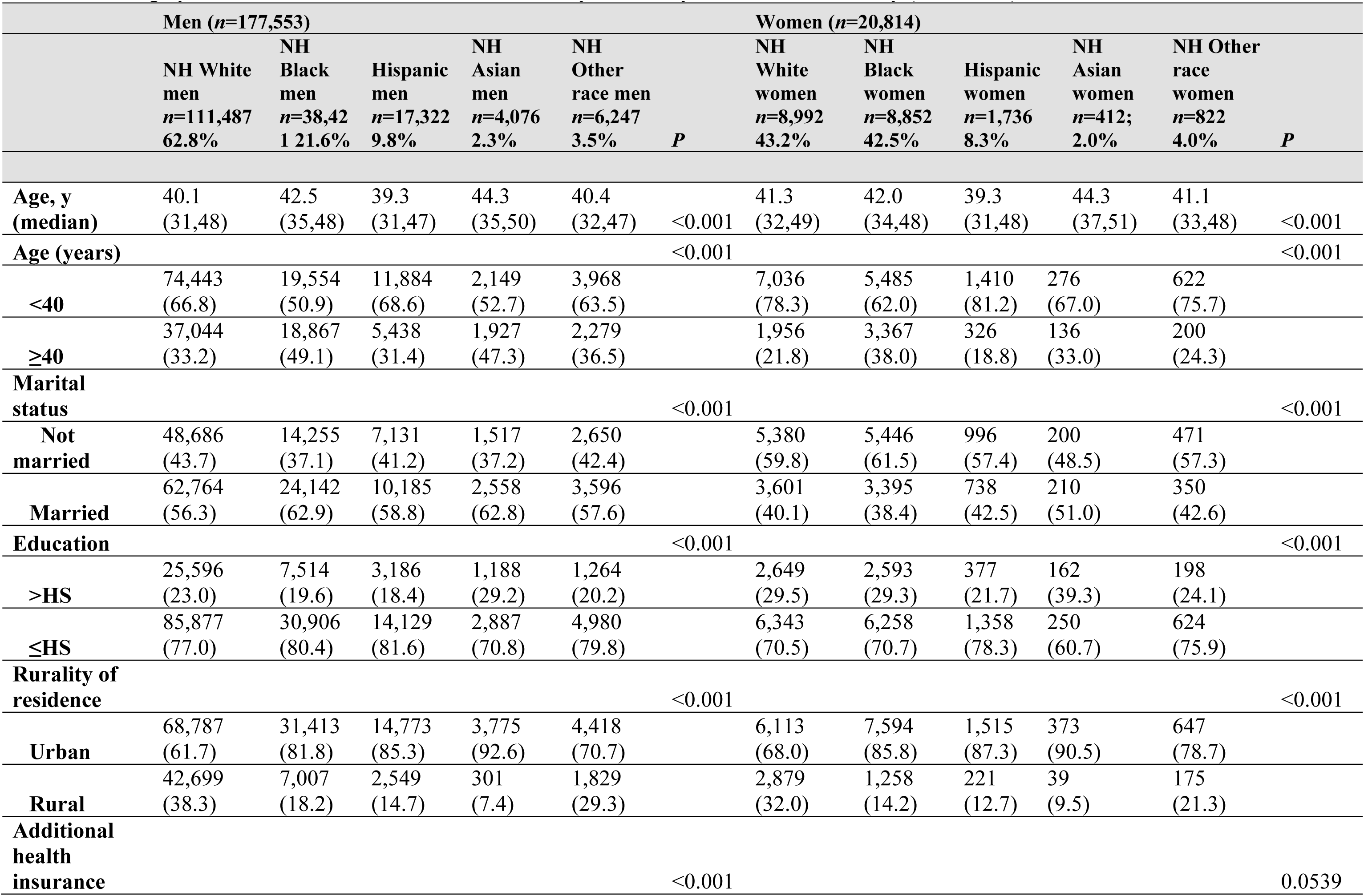
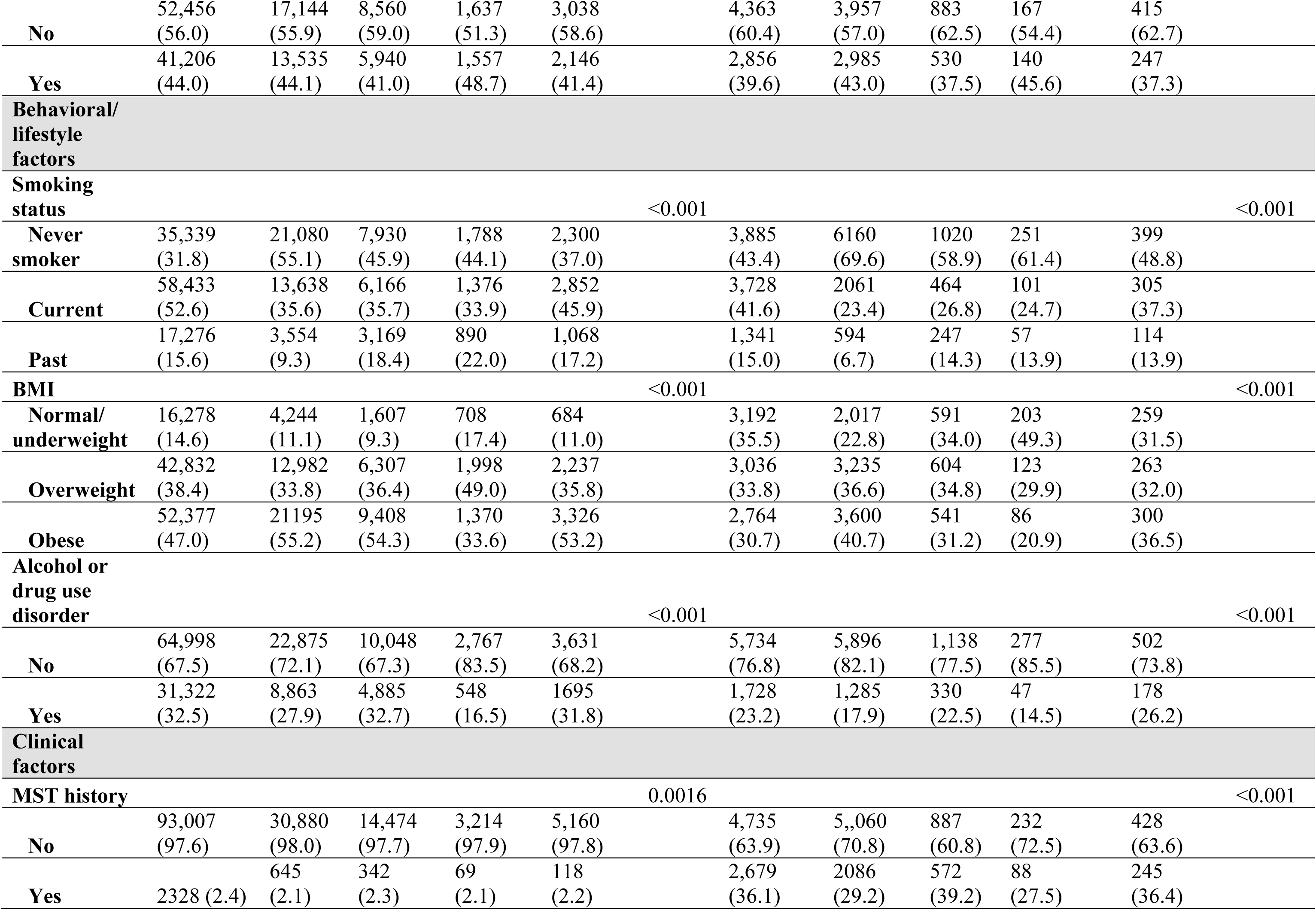
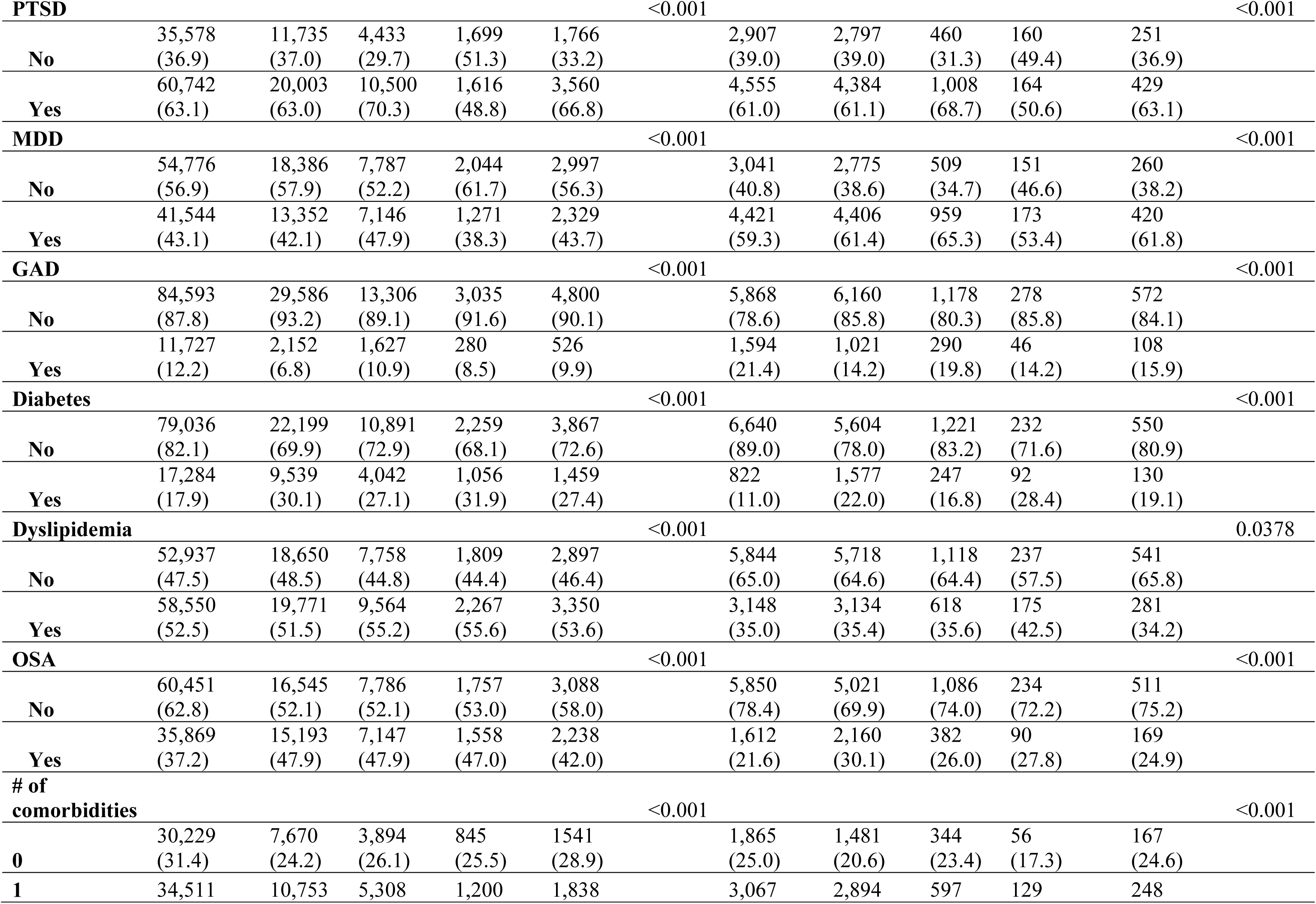
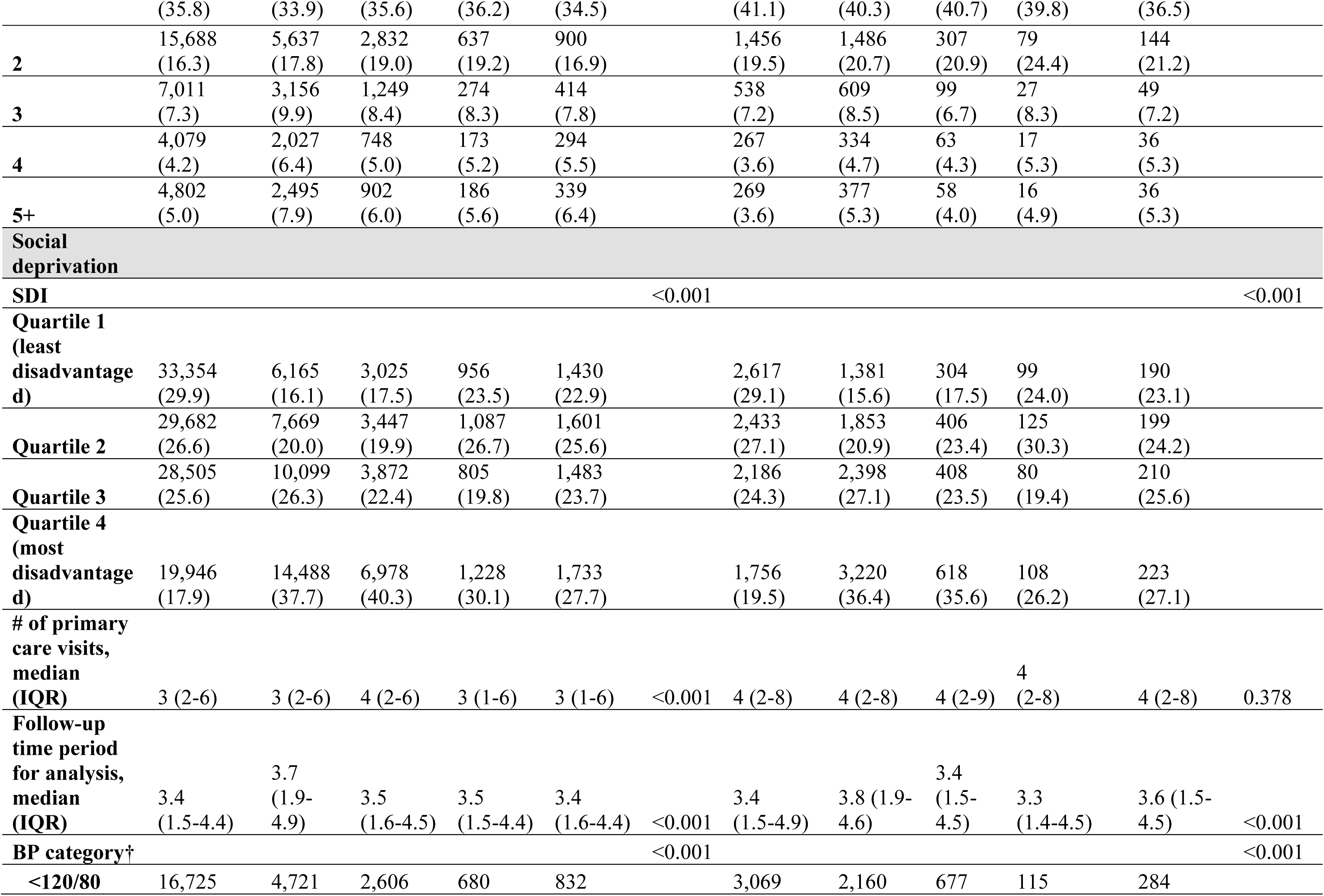
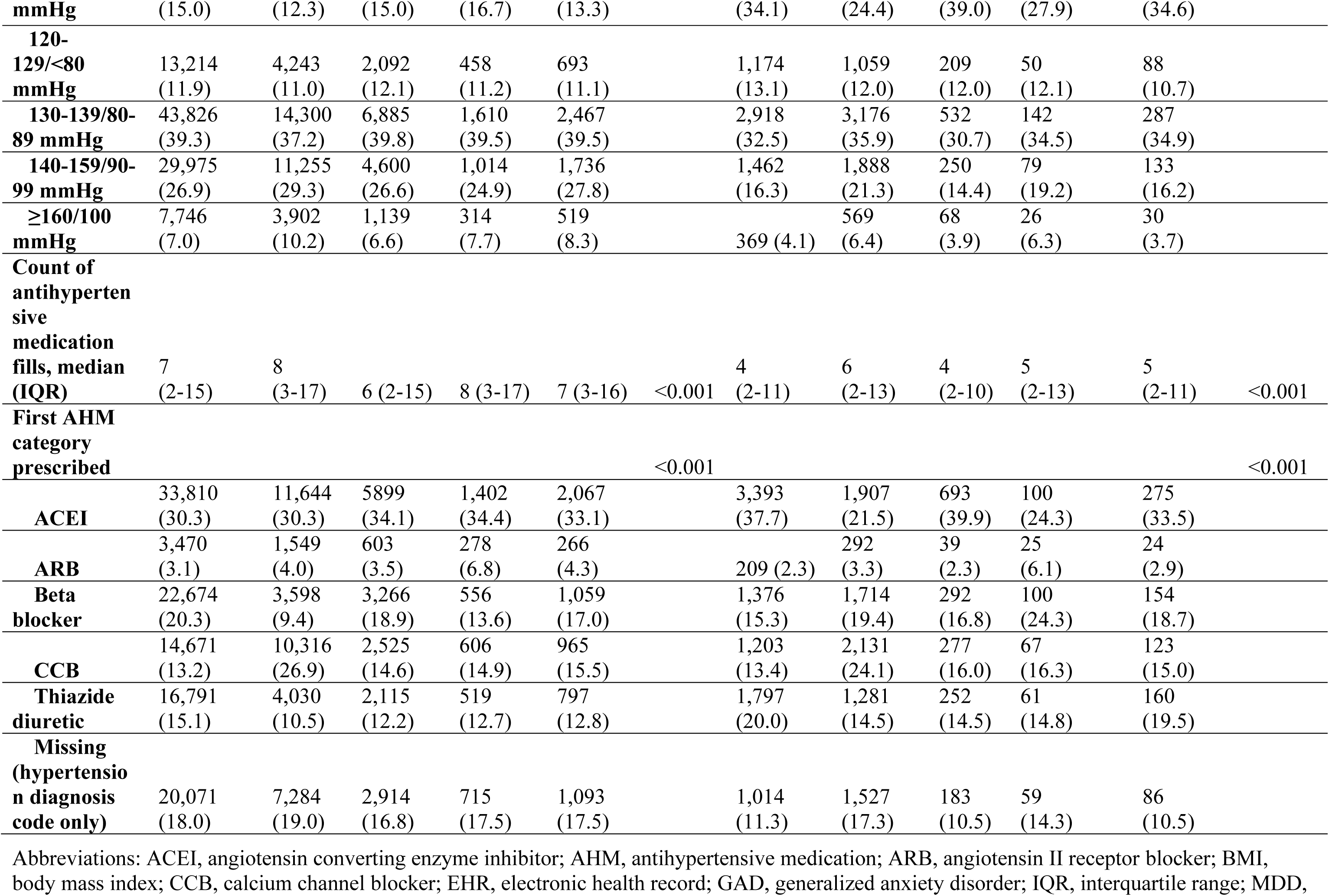
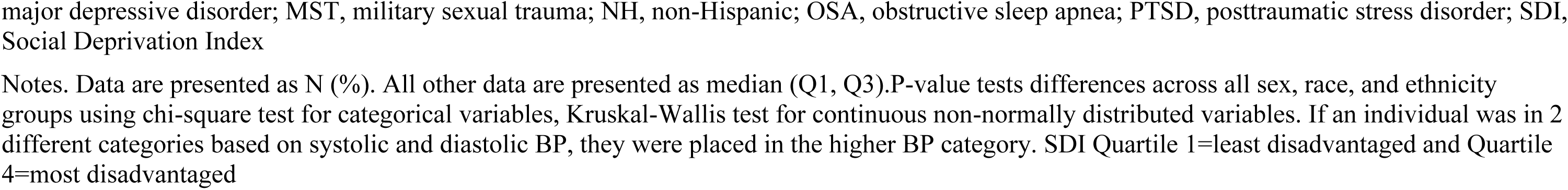
Demographic, behavioral, and clinical characteristics, presented by sex, race, and ethnicity (n=198,367).

When examining the sample of women with hypertension, there was a similar age distribution between White, Hispanic, and Other subgroups, although a greater proportion of Black and Asian women were older than 40. In each subgroup more women had a high school education or less and lived in an urban vs. rural locale. Compared to other subgroups, White women were more likely to currently smoke and to have an anxiety diagnosis but were less likely to be diagnosed with diabetes. In contrast, women who identified as Black, and Hispanic, and Other race showed the highest prevalence of PTSD and depression among the groups. For about 1 in 3 women from each group, baseline BP was in the range of 130-139/80-80mmHg. All groups of women were typically prescribed an ACE-inhibitor first, except for Black women who were more likely to be prescribed a calcium channel blocker.

As depicted in Table 4, within one year of meeting criteria for hypertension, 55% of men and 69% of women had controlled BP. Men who were ≥40 years, Asian or Hispanic, married, with more than a high school education, insurance, no history of substance use disorder, and a normal/underweight BMI, had significantly better BP control than men in comparison groups. Among women, a greater proportion with controlled BP were <40 years old, White, or Hispanic. Two years after the onset of hypertension, approximately 54% of men and 67% of women had controlled BP. For men, BP control was again better for those who were younger, married, more educated, and without substance use disorders or PTSD. Among women, BP control was significantly better for those who were younger, identified as White or Hispanic, and without a substance use history than women in comparison groups. At five years beyond initial hypertension documentation, 53% of men and 66% of women demonstrated BP control. Among men, those who were younger, Asian, married, educated, without a substance use disorder history, and without an OSA diagnosis showed better BP control than men in reference groups. Similarly, women who were younger, White or Hispanic, and in the lowest SDI quartile showed significantly better BP control compared to women in reference groups.

**Table 4.**
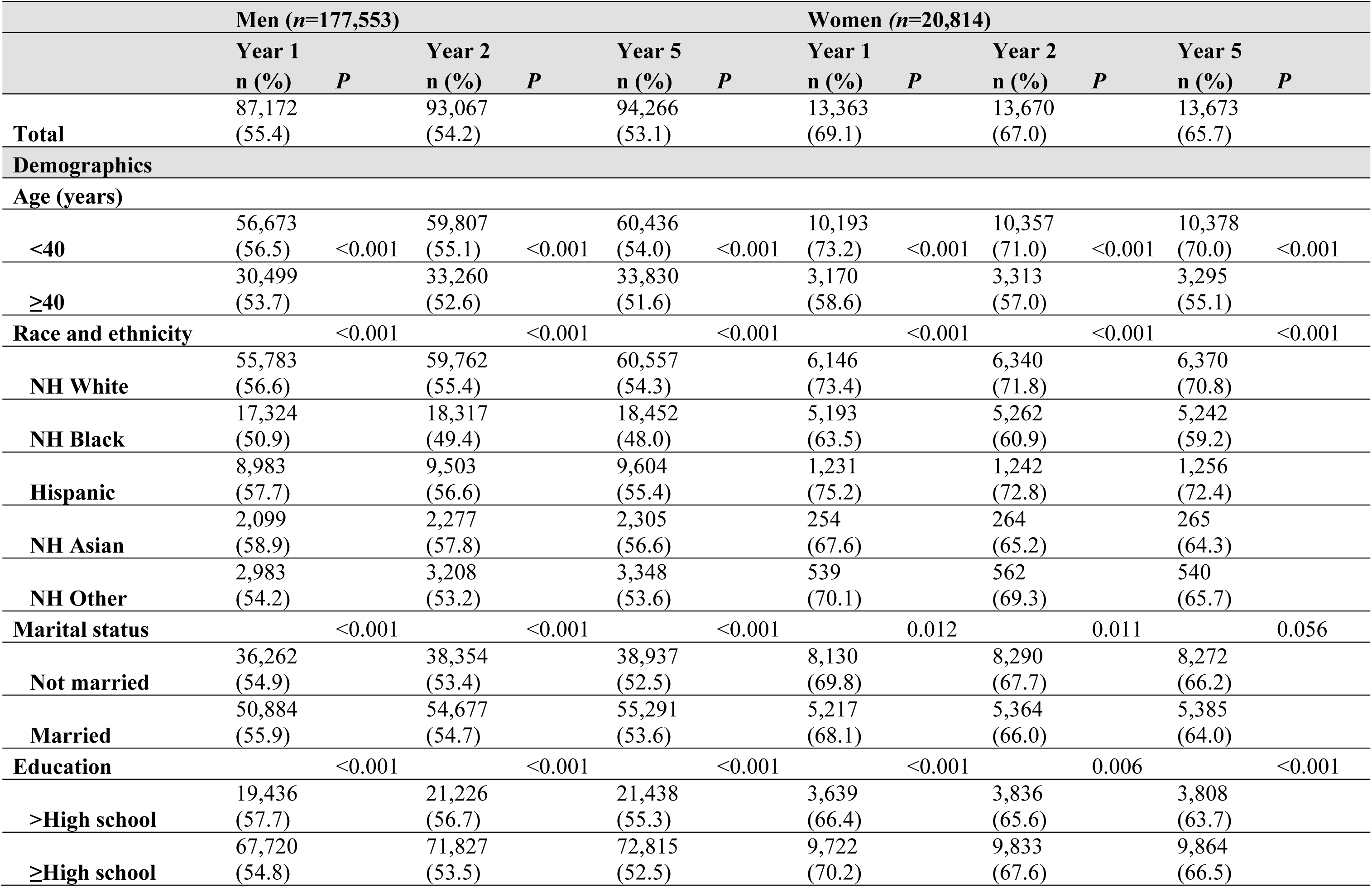
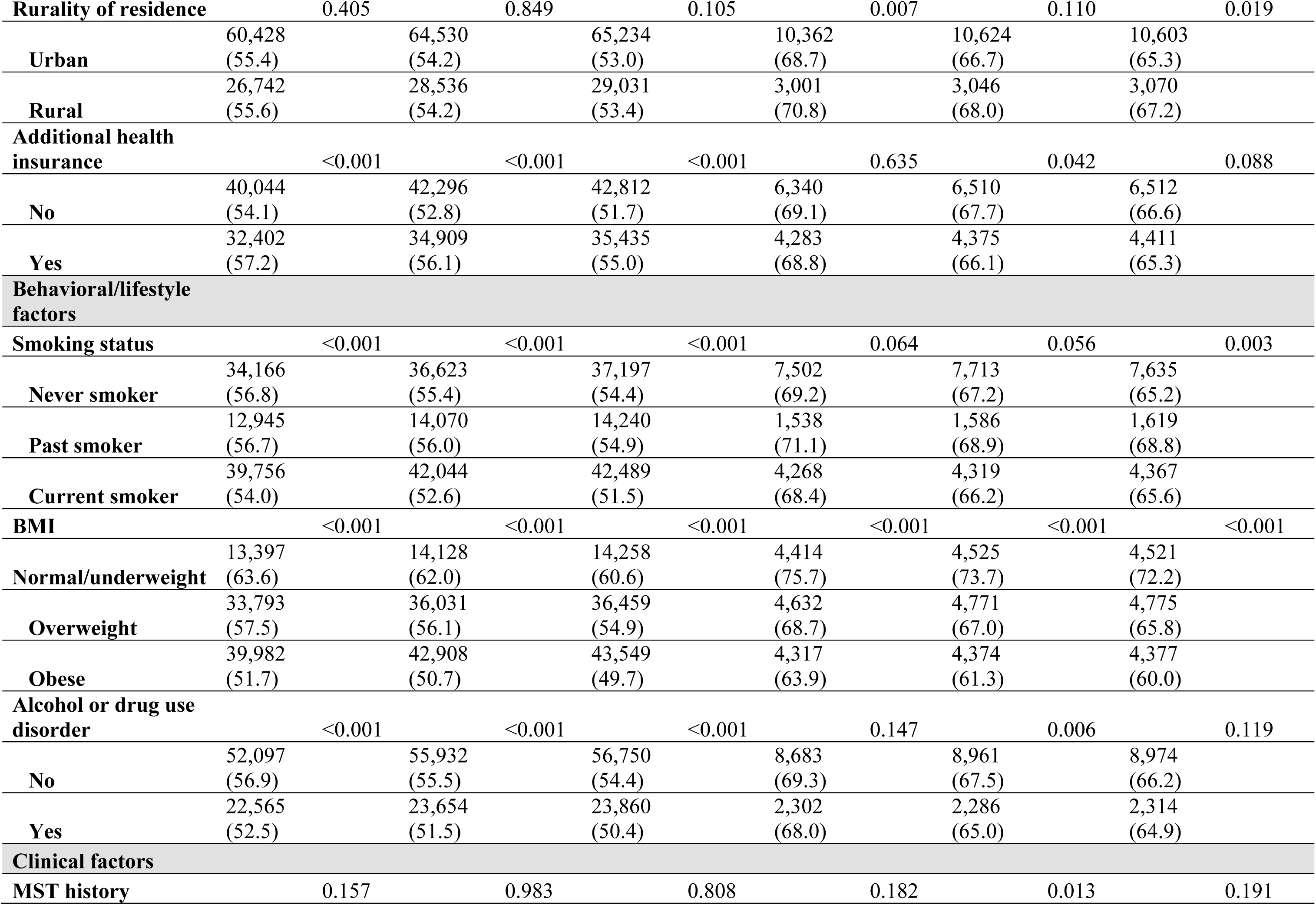
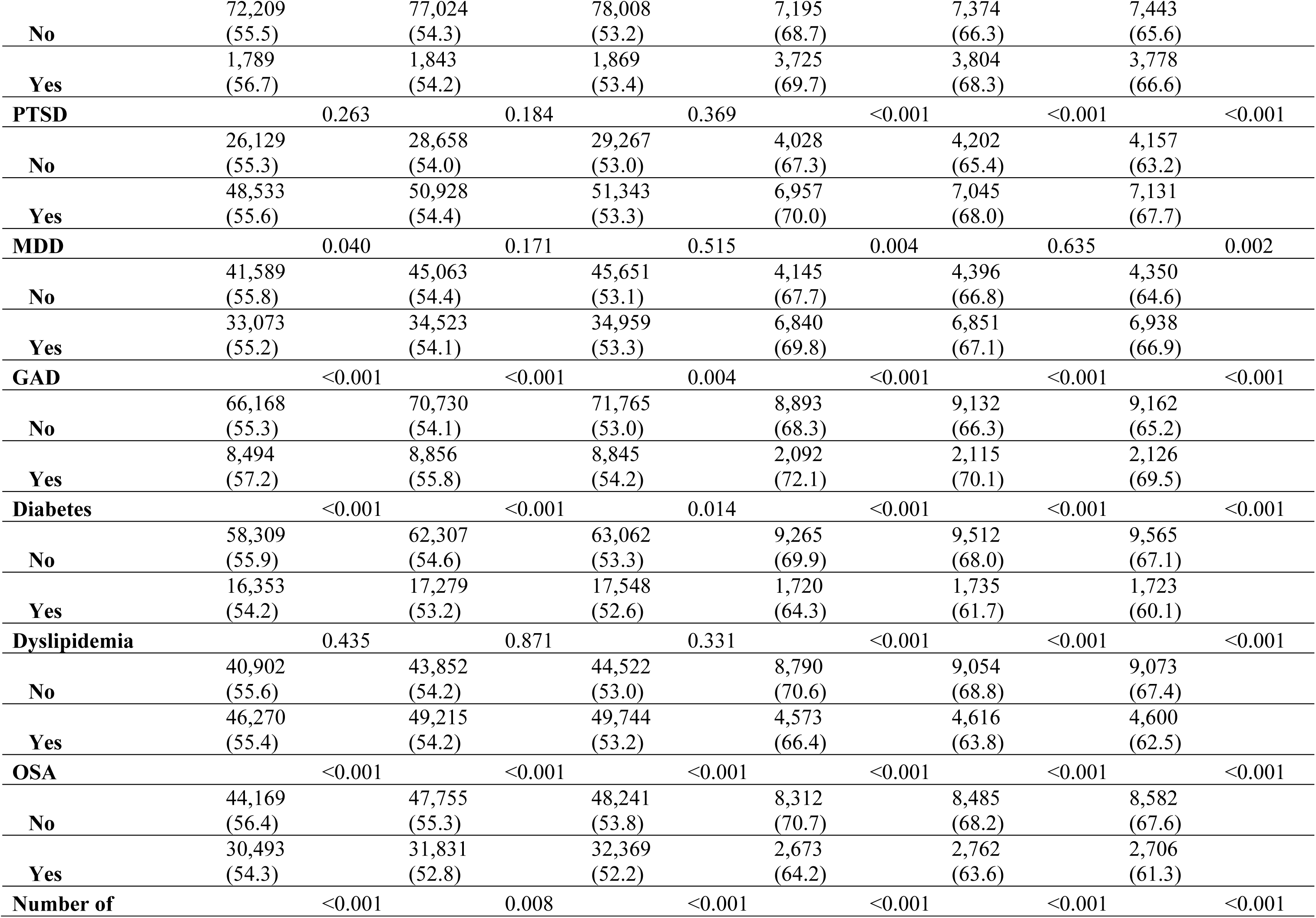
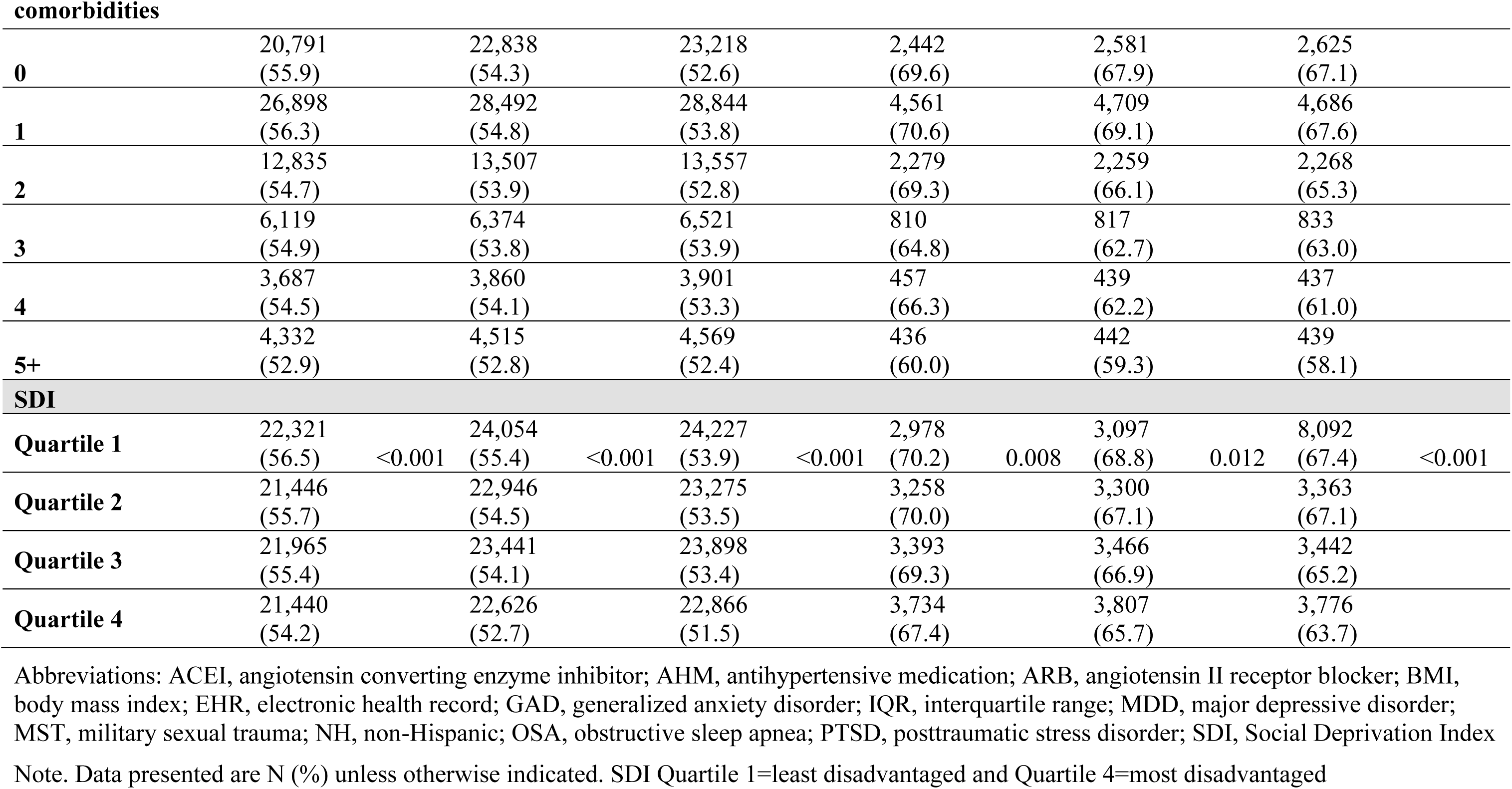
Proportion of the cohort with controlled BP at years 1, 2, 5 after a hypertension diagnosis, by descriptive and clinical factors, n=198,367.

In unadjusted models of BP control at year 1, women had a 90% greater likelihood of BP control than men. Results from the adjusted multivariable tests of Years 1, 2, and 5 are depicted in Table 5. After multivariable adjustment for demographics, behavioral factors, and clinical factors, women still showed a 66% greater odds of BP control than men (95% CI: 1.59-1.63). Younger age, Hispanic ethnicity, and having a diagnosis of PTSD, were associated with increased odds of controlled BP at 1 year after meeting criteria for hypertension. Yet, Black, Asian, and Other racial groups, being unmarried, a current smoker, overweight or obese, and a history of substance abuse were associated with a lower likelihood of controlled BP. Finally, the interaction between sex and race/ethnicity was significant (*p*=0.006). Compared to White men, both Black (OR: 0.81 [95% CI: 0.78-0.84]) and Other-identified groups of men showed a significantly lower likelihood of BP control (OR: 0.88 [95% CI: 0.81-0.95]). Similarly, compared to White women, the likelihood of BP control was lower for women who identified as Black (OR: 0.67 [95% CI: 0.61-0.74]), Asian (OR: 0.65 [95% CI: 0.46-0.90]), and women who were members of Other racial groups (OR: 0.76 [95% CI: 0.60-0.96]).

**Table 5.** Multivariable Models for the Effects of Sex on Odds of BP Control at 1, 2, and 5 Years.

|  | Year 1 |  | Year 2 |  | Year 5 |  |
| --- | --- | --- | --- | --- | --- | --- |
|  | OR<br>(95% CI) | *P* | OR<br>(95% CI) | *P* | OR<br>(95% CI) | *P* |
| Women | 1.66<br>(1.59-1.73) | <0.001 | 1.62<br>(1.54-1.71) | <0.001 | 1.60<br>(1.53-1.69) | <0.001 |
| **Demographics** |  |  |  |  |  |  |
| <40 years<br>(vs ≥40 years) | 1.24<br>(1.20-1.28) | <0.001 | 1.22<br>(1.19-1.26) | <0.001 | 1.20<br>(1.16-1.23) | <0.001 |
| Black (vs White) | 0.78<br>(0.75-0.81) | <0.001 | 0.76<br>(0.74-0.79) | <0.001 | 0.78<br>(0.75-0.80) | <0.001 |
| Hispanic (vs White) | 1.09<br>(1.04-1.14) | <0.001 | 1.08<br>(1.03-1.13) | <0.001 | 1.04<br>(0.99-1.09) | 0.093 |
| Asian (vs White) | 0.90<br>(0.81-0.99) | 0.038 | 0.99<br>(0.90-1.09) | 0.770 | 0.98<br>(0.89-1.07) | 0.621 |
| Other (vs White) | 0.87<br>(0.80-0.93) | <0.001 | 0.89<br>(0.83-0.96) | 0.002 | 0.93<br>(0.87-1.00) | 0.039 |
| Not married (vs married) | 0.92<br>(0.90-0.95) | <0.001 | 0.92<br>(0.89-0.94) | <0.001 | 0.93<br>(0.91-0.96) | <0.001 |
| **Behavioral/lifestyle factors** |  |  |  |  |  |  |
| Past smoker (vs never) | 0.96<br>(0.92-1.00) | 0.073 | 0.98<br>(0.94-1.02) | 0.409 | 1.00<br>(0.96-1.04) | 0.925 |
| Current smoker (vs never) | 0.88<br>(0.85-0.91) | <0.001 | 0.88<br>(0.85-0.91) | <0.001 | 0.88<br>(0.85-0.90) | <0.001 |
| Overweight (vs normal/under) | 0.73<br>(0.70-0.76) | <0.001 | 0.75<br>(0.73-0.79) | <0.001 | 0.76<br>(0.73-0.79) | <0.001 |
| Obese (vs normal/under) | 0.56<br>(0.54-0.58) | <0.001 | 0.57<br>(0.55-0.60) | <0.001 | 0.59<br>(0.56-0.61) | <0.001 |
| Alcohol abuse or drug use (vs never) | 0.79<br>(0.77-0.82) | <0.001 | 0.80<br>(0.78-0.83) | <0.001 | 0.81<br>(0.78-0.83) | <0.001 |
| **Clinical risk factors** |  |  |  |  |  |  |
| MST ever (vs no history) | 1.05<br>(0.98-1.13) | 0.135 | 1.04<br>(0.97-1.10) | 0.297 | 0.99<br>(0.93-1.05) | 0.685 |
| PTSD ever (vs never) | 1.07 | <0.001 | 1.07 | <0.001 | 1.06 | <0.001 |

|  | (1.04-1.11) |  | (1.04-1.10) |  | (1.03-1.09) |  |
| --- | --- | --- | --- | --- | --- | --- |
| # of comorbidities | 1.00<br>(0.99-1.01) | 0.412 | 1.01<br>(1.00-1.01) | 0.237 | 1.01<br>(1.00-1.02) | 0.005 |
| OSA ever (vs never) | 1.00<br>(1.00-1.00) | 0.197 | 1.00<br>(1.00-1.01) | 0.054 | 1.01<br>(1.00-1.01) | 0.058 |
| Diabetes (vs none) | 0.98<br>(0.95-1.01) | 0.153 | 0.94<br>(0.92-0.97) | 0.282 | 0.97<br>(0.95-1.00) | 0.581 |
| Dyslipidemia (vs none) | 1.03<br>(0.99-1.07) | 0.160 | 1.02<br>(0.98-1.06) | 0.563 | 1.01<br>(0.97-1.05) | 0.282 |
| **Primary care utilization** |  |  |  |  |  |  |
| # of visits in the first 2 years | 1.02<br>(0.99-1.05) | 0.312 | 1.01<br>(0.98-1.04) | 0.023 | 1.02<br>(0.99-1.05) | <0.001 |

Abbreviations: CI, confidence interval; MST, military sexual trauma; OR, odds ratio;
OSA, obstructive sleep apnea; PTSD, posttraumatic stress disorder; SDI, Social Deprivation Index

At Year 2, women had an 83% greater likelihood of BP control than men, an association that decreased to 62% in the fully adjusted model (95% CI: 1.54-171). Younger age and greater primary care utilization were also associated with a greater odds of BP control. Black and Other races, being unmarried, current smoking, overweight or obesity, and substance use were associated with the likelihood of controlled BP. The Sex*Race/Ethnicity interaction was again significant at this timepoint (*p*<0.001), with other patterns similar to those observed in Year 1.

Finally, at Year 5 women showed an 80% greater, unadjusted odds of BP control than men. After adjustment, women showed a 60% greater likelihood of BP control vs. men (95% CI: 1.53-1.69). As observed with earlier timepoints, younger age, a PTSD diagnosis, and primary care utilization were each associated with BP control. Conversely, patients who were Black and Other race, unmarried, current smokers, overweight or obese and with a substance use history had lower BP control. The Sex*Race interaction was again significant (*p*<0.001), with other associations like those observed in Years 2 and 5.

Finally, SDI was introduced in exploratory analyses to determine if social deprivation would attenuate effects of sex, race, or ethnicity on BP control. Overall, at Year 1, patients who resided in the most disadvantaged areas showed a 5% lower odds of BP control than the least disadvantaged group (Quartile 4 vs. Quartile 1; 95% CI: 0.91-0.99). This association remained in Year 2 (OR 0.94 [95% CI: 0.91-0.98]) and Year 5 (OR 0.95 [95% CI: 0.91-0.99]). Still, the previous associations of sex, race, and ethnicity and likelihood of BP control were maintained each year. When stratifying the SDI analyses by demographic subgroups, the effect of SDI on BP control only remained for select groups. Among men at Year 1, those who identified as Black and belonged to most disadvantaged SDI group (Quartile 4) had an 11% lower likelihood of BP control than those in the most advantaged group (Quartile 1; 95% CI 0.81-0.99). This association was maintained at the Year 2 (OR 0.89 [95% CI: 0.81-0.98]) and Year 5 timepoints (OR 0.88 [95% CI: 0.80-0.97]). Men who identified as Other and in SDI Quartile 2 also had a 23% lower odds of BP control than those in Quartile 1 (95% CI: 0.61-0.96). Finally, Black women in SDI Quartile 3 showed a 21% lower likelihood of BP control at the Year 2 and 5 timepoints (both 95% CI: 0.61-0.96).

## DISCUSSION

In this 16-year, prospective, nationwide cohort study of younger adults, 17% of patients met criteria for hypertension. One year after meeting initial criteria for hypertension, 45% of men and 31% of women had not achieved BP control. After adjustment for other demographic, behavioral, and clinical variables, within one year, BP control was 66% more likely for women than men, and this difference was consistently observed up to five years later. Yet contrary to our hypothesis, the proportions of men and women who achieved BP control *decreased* across the 2- and 5-year timepoints, rather than increasing. In addition to differences by sex, patients who were younger and with a PTSD diagnosis were more likely to have BP control while those who were older, unmarried, current smokers, obese, and with a substance use disorder were all less likely to demonstrate BP control, factors that were similar for men and women, and over time.

The sex-specific prevalence of BP control also differed by race and ethnicity. Among men and women, adults who identified as Black and Other race showed worse BP control than their White counterparts. However, women who identified as Asian also had significantly worse BP control than patients who were same-sex and White. Altogether, these data show that disaggregating the population of patients with hypertension by sex, race, and ethnicity is important to determine which subgroups may require more clinical and social support to achieve BP control.

The present study revealed differences between men and women in both the identification of hypertension and in subsequent BP control. Per 2017-2018 data from the U.S. Centers for Disease Control and the National Health and Nutrition Examination Survey (NHANES), the prevalence of hypertension among younger and middle-aged adults is 31%-59% for men and 13%-50% for women.^31^ Despite patients’ greater risk for later CVD,^1–5^ in the present cohort, considerably fewer adults met criteria for hypertension than expected – 18% of men and 15% of women. These differences may be due to a lingering healthy soldier effect in the initial years following service,^32^ which has been estimated to lower health-related risk by 10%-25%,^33^ to underdiagnosis of hypertension among this complex patient population, and to slightly different inclusion criteria as NHANES participants can also meet criteria for hypertension based on BP readings. To conservatively estimate the prevalence of hypertension in the present study, and given greater multimorbidity observed among Veterans, BP was not used as a sole inclusion criterion. Variability in hypertension documentation was also observed. One-third of adults in VA care were prescribed an AHM but did not have a documented diagnosis of hypertension. As hypothesized, significantly fewer men than women – 31% versus 49% – were missing a hypertension diagnosis. Contrary to expectation, a higher percentage of Black men and women were each more likely to meet both hypertension criteria than same-sex individuals who were characterized as other race and ethnicity groups. Perhaps those without medications are less aware of their hypertension status, or among those without a documented hypertension diagnosis, clinicians may be less attentive to BP control, both questions that could be pursued in future investigations.

Characterizing BP control by sex is important in light of the growing evidence that men and women differ in associations from BP to CVD outcomes.^34^ While there are known sex differences in the receipt of diagnosis and guideline-directed management of established CVD among Veterans served by the VA,^35^ the degree to which BP control differs by sex is relatively unexplored among younger Veterans, for whom early BP control can have the greatest effect for later morbidity and mortality. Within one year of meeting study criteria for hypertension, 55% of men and 69% of women had controlled BP, similar that were similar to NHANES data from the general population.^25,36^ Confirming our hypothesis, women were 66% more likely to achieve initial BP control than men, and maintained this greater likelihood of BP control for five years.

In contrast, NHANES data showed little difference in hypertension control rates by sex.^8^ Other, past investigations of sex differences in BP control yielded conflicting results, with some showing that men or women each have better BP control than adults of a comparable age, or that there is no difference by sex, with variability in these results based on age, race, and ethnicity.^16,25,37^ Sex differences in hypertension management could begin with the patient’s knowledge about their health. Compared to younger men, same-aged women are 35% more likely to be aware of their hypertension,^38^ a difference that may be attributable to women’s greater healthcare utilization, and may explain results for the present study. Among Veterans specifically, younger women utilize VA care more frequently than their male counterparts,^29^ providing more opportunities for BP monitoring and management. In consideration of these converging lines of evidence, sex-specific approaches to BP management may be helpful, but have not yet been developed and tested.^39,40^

Upon examining sex, race, and ethnicity concurrently, as hypothesized, a greater proportion of White adults had BP control than Blacks at each timepoint. White and Black women also each showed significantly better BP control than their same-race male counterparts, associations that persisted through five years. When comparing BP control within sex, Black men and women each had worse BP control than same-sex individuals who identified as White. Thus, the present findings provide two important extensions of past data among Veterans^41–43^ – examining sex by race/ethnicity interactions and comparing racial and ethnic subgroups within men and women, respectively. In the present results, our study replicates some of the racial disparities observed in NHANES and other large datasets.^8^ Given that the VA system is designed to offer uniform care to all Veterans, disparities in the outcomes of treatment may occur due to clinician judgment and bias, and the quality of care at facilities more frequented by racial and ethnic minorities groups.^44,45^ It is also possible that subgroup differences in BP control are due to racial differences in trust, awareness, adherence, and health beliefs,^9^ which should also be investigated among younger Veterans, particularly women.^45^

SDoH can influence vulnerability to hypertension, access to treatment, and long-term BP management,^17^ particularly among women.^46^ Yet, adding the SDI as a proxy of SDoH did not significantly alter associations with BP control. Overall, patients who were categorized as most disadvantaged showed only a 5% lower likelihood of BP control compared to the least disadvantaged group. However, in adjusted analyses stratified by demographic subgroups, effects of SDI were specific to non-Hispanic Black adults only, particularly men. The VA is the largest healthcare system in the U.S. and is designed to provide a uniform standard of care to Veterans that supersedes social disadvantages, which could explain why inclusion of SDI did not greatly influence the likelihood of BP control by sex, race, or ethnicity. Still, Black patients with hypertension and who show the greatest social deprivation also have the greatest difficulty with BP control. Among Veterans receiving VA care, SDoH-related disparities may be mostly encapsulated by sex, race, and ethnicity, as in these analyses, then by other variables. Another recent retrospective cohort study of Veterans with heart failure found no meaningful differences in treatment receipt between those who lived in rural or urban locations.^47^ However, as SDI was a composite index, it is worth investigating if specific social deprivation variables – e.g., unemployment, living conditions – are more predictive of BP control among younger Veterans, and if associations are specific to certain subgroups or vary by disadvantage level.

This research improves our knowledge of VA-wide efforts to manage BP among younger Veterans, potential inequities in clinical care for hypertension, and the effect of these two major events on cardiovascular risk management among younger Veterans. In support of the observed disparities in the provision of AHM, among a sample of adults with similar BP, women were less likely to receive AHM at all.^48^ In terms of first-line therapy for reproductive-age women, when AHM are prescribed, a lower percentage of women were prescribed certain medications than men (i.e., ACE inhibitors and angiotensin receptor blockers [ARBs]), due to potential for teratogenicity among women of reproductive age,^49^ but these medications were still prescribed to over 20% of women, suggesting a lower quality of care when managing BP. Overall, sparse data indicate that women respond better to diuretics, ACE inhibitors and beta-blockers,^50^ but those studies were based on other samples and there is little evidence concerning the comparative effectiveness of AHM in younger women. Improving BP control among young and middle-aged adults requires improved implementation of the existing guidelines – appropriate measurement of BP during office visits, and tools for home BP measurement, special consideration for additional changes needed by those with a history of uncontrolled hypertension, or with a past hypertensive disorder of pregnancy or during birth – including earlier encouragement of proactive lifestyle interventions and pursuit of pharmacologic options.^51^ Investigating the processes of BP assessment in the VA, such as alignment with BP management guidelines in the first year of care or the frequency with which home BP monitoring is used to verify BP assessed in clinic,^26^ will be valuable to determine how best to help younger men and women Veterans with hypertension and uncontrolled BP in the first year, and then over time.

Strengths of this investigation include accounting for hypertension among all men and women post-9/11 Veterans receiving VA care, and across a follow-up period of 16 years. The characteristics of this cohort also offered a singular opportunity to examine the specific effects of sex, race, and ethnicity, and multiple risk factors for hypertension and CVD, on BP control.

Furthermore, hypertension was rigorously defined based on two mutually inclusive criteria – EHR diagnosis and AHM use. Finally, by limiting the sample to those with a documented zip code, it was possible to explore if demographics or SDoH were the most influential predictors of BP control.

The limitations of this investigation should also be enumerated. First, as the sample consisted of only post-9/11 Veterans who sought VA care, the results may not be generalizable to former military servicemembers from other eras, men and women who do not receive care through the VA, or to adults in the general population. Second, as this was a prospective cohort study involving EHR data, there is a risk for misclassification bias, selection bias, and missing data.

For example, hypertension is also likely underdiagnosed, including among younger Veterans and those who are women, and to ensure our estimates were conservation we did not include patients based on BP readings from the EHR. Third, it was not possible to control for all aspects of lifestyle and behavioral factors or medication adherence in this investigation. Future investigations are required to better account for physical activity, diet, sleep duration, and psychosocial stress, particularly as those factors show differential associations by sex, race, ethnicity, and SDoH. Fourth, clinic BP measurement does not always reflect BP patterns indexed in non-clinical settings. Ambulatory BP monitoring or home BP monitoring are recommended to confirm a diagnosis of hypertension and monitoring BP control but was not used in these data.^26,30^ Fifth, we did not examine demographic differences in BP control by type of AHM used or among those taking monotherapy vs combination therapy, which was beyond the scope of the present analyses and will be a valuable future investigation.^52^ Sixth, patients could have received care outside of the VA, including management of hypertension. Relatedly, exploring differences in BP assessment and management by these groups is a vital next step.

### Perspectives

BP control is a crucial aspect of CVD risk management. In this cohort of younger and middle-aged Veterans receiving care through the VA, over half of patients demonstrated BP control in the first year after meeting criteria for hypertension, patterns that persisted through two and five years later. At each timepoint, women and White, non-Hispanic patients were more likely to show BP control than men and patients who identified as non-White, indicating that BP control warrants greater focus for male and Black Veterans. Relatively stable rates of BP control over the subsequent five years suggests that the first year of hypertension care is crucial for all.

Finally, social deprivation did not significantly alter these associations, suggesting that the VA system may protect against some of the social inequity that is observed in the U.S. general population. Improving the identification of hypertension and expeditious, initial BP control in younger men and women should be a priority for VA and non-VA providers alike.

### Novelty and Relevance

- What Is New?

1. This is the largest prospective cohort study of younger men and women Veterans with a focus on determining the prevalence of hypertension and individual differences in blood pressure (BP) control.
2. Unlike many previous examinations of BP control in younger adults and Veterans, data were based on electronic health records rather than self-report of hypertension and related BP control.
3. Following the cohort up to 5 years after they met criteria for hypertension allowed us to determine how BP control may change over time, and to identify individual differences in patterns of BP control, including exploring the role of social deprivation.
- What Is Relevant?

1. More men than women had both documentation of a hypertension diagnosis and an antihypertensive prescription (51% vs. 38%). By race and ethnicity, a greater proportion of Black men and women had both a hypertension diagnosis and an antihypertensive prescription and members of other race and ethnic groups.
2. Within one year of meeting criteria for hypertension, about 1 in 2 men and 2 of 3 women had controlled BP, but these proportions decreased slightly over the next 5 years. Non-Hispanic, White patients had the highest rates of BP control at each timepoint, while Black adults and Asian women had the lowest BP control at 1 year. This distinction persisted over time for Black patients.
3. Patients with the greatest social deprivation showed a 5% lower likelihood of BP control across 5 years, and Black men and women with greater social deprivation had the lowest odds of BP control.
- Clinical/Pathophysiological Implications?

1. Closely monitoring BP control for the first year should be sufficient for most patients with hypertension, but it is likely that regular, intensive BP management should extend through 5 years and beyond.
2. Younger patients who are Black, and Asian women, may especially benefit from additional BP management in the first year, and certain disadvantaged groups likely require greater, regular support to achieve BP control over time.

## Data Availability

Data used for these analyses are covered under a proprietary data use agreement with the Department of Veterans Affairs and are not available for distribution by the authors.

## Acknowledgements

We would like to thank all the Veterans for their service and contributions to this investigation. The views and opinions of authors expressed herein do not necessarily state or reflect those of the United States Government.

## Non-standard Abbreviations and Acronyms

ACE-inhibitor: Angiotensin-converting enzyme inhibitors
AHM: antihypertensive medication
ARBs: angiotensin receptor blockers
BMI: body mass index
BP: blood pressure
CI: confidence interval
CVD: cardiovascular disease
EHR: electronic health record
ICD-9-CM and 10: International Classification of Diseases, Ninth Revision, Clinical Modification and Tenth Revision
MST: military sexual trauma
NHANES: National Health and Nutrition Examination Survey OR, odds ratio
OSA: obstructive sleep apnea
PTSD: posttraumatic stress disorder
SDI: Social Deprivation Index
SDoH: social determinants of health
VA: Department of Veterans Affairs

